# Increased amygdala volume and functional connectivity with cognitive control networks in chronic migraine

**DOI:** 10.1101/2020.08.31.20185397

**Authors:** Danielle D. DeSouza, Samuel R. Krimmel, Bharati M. Sanjanwala, Addie Peretz, Vinod Menon, David A. Seminowicz, Robert P. Cowan

## Abstract

**Objective:** To characterize the role of the amygdala in episodic (EM) and chronic (CM) migraine, we evaluated amygdala volumes, functional connectivity (FC), and associations with clinical and affective measures.

**Methods:** Eighty-eight patients (44 with EM and 44 age- and sex-matched patients with CM) completed anatomical and resting-state functional MRI scans. Amygdala volumes and resting-state FC to three core large-scale cognitive control networks (default mode (DMN), salience (SN), central executive (CEN)) were compared between groups. Associations between amygdala volume and FC, measures of headache severity (frequency and intensity), and cognitive-affective measures (depression, anxiety, pain catastrophizing) were evaluated.

**Results:** Compared to EM, patients with CM had larger amygdala volume bilaterally. Headache frequency and intensity were associated with increased left and right amygdala volume, and depression was associated with increased right amygdala volume. Patients with CM also demonstrated increased left amygdala FC with the DMN, which across patients was related to headache frequency. Left amygdala FC to the SN was correlated with headache intensity while right amygdala FC to the CEN was correlated with pain catastrophizing.

**Conclusion:** Our findings reveal increased amygdala volume and FC with large-scale neurocognitive networks in patients with CM compared to EM. Aberrant amygdala volume and FC measures were associated with increased migraine severity, depression, and pain catastrophizing, pointing to a link between emotion and pain in migraine. Our findings provide novel insights into amygdala involvement in chronic migraine and may inform future interventions aimed at preventing the progression of both headache and its negative cognitive-affective symptoms.

## INTRODUCTION

Episodic (EM) and chronic (CM) migraine are part of a spectrum of migraine disorders, however, they are viewed as separate clinical entities based on the frequency and duration of headache symptoms^1^. One important clinical problem is the progression of EM (<15 headache days/month) to CM (≥ 15 headache days/month with migraine features on ≥ 8 days/month for ≥ 3 months), a more disabling condition associated with greater migraine-related disability, poorer quality of life, and increased psychiatric comorbidities^2–5^. Migraine progression, or chronification, occurs at a rate of 3% each year^6,7^. While there is evidence to support persistent alterations across sensory, autonomic, and affective circuits^8^, the pathophysiological mechanisms underlying migraine chronification are not fully understood.

One brain structure of growing interest for its key roles in sensory integration, nociceptive processing, emotional responses, and affective states^9^ is the amygdala^9,10^. Mounting support for amygdala plasticity underlying sustained pain has been provided by preclinical models^8,11,12^ and functional MRI studies of clinical of pain^13,14^. While the literature supports a role for altered amygdala structure and/or function in migraine^15–21^, few studies have directly compared EM to CM^16,17,20^. Additionally, mixed methods limit the ability to compare results across the migraine literature, especially to recent studies showing large-scale brain network differences^22–25^.

Here, we compared amygdala volumes and functional connectivity to three large-scale brain networks^26^ in EM and CM. Based on previous research^16,17,20^, we hypothesized larger amygdala volumes and increased amygdala-network connectivity in CM, and positive associations with headache and cognitive-affective measures.

## METHODS

### Participants

Participants were recruited as part of two larger studies acquiring clinical, behavioral, and MRI data to examine biomarkers of chronic daily headache (patients with CM, Stanford University) and mindfulness based stress reduction for migraine (patients with EM, University of Maryland, Baltimore and Johns Hopkins University)^27^. Prior publications including these data^27,28^ did not investigate amygdala structure and/or function between EM and CM in relation to headache and cognitive-affective measures. Patients were eligible for the current study if they were at least 18 years of age and met the diagnostic criteria for EM or CM with or without aura for at least three months according to the International Classification of Headache Disorders (ICHD)-3^1^, as determined by a physician. Patients with CM were recruited between January 2015 and November 2017 for the CM group and June 2014 to February 2017 for the EM group. Participants were excluded if they had MRI contraindications, elected not to participate in the MRI scanning session, or had a history of severe neurologic or psychiatric comorbidities. Neither group was asked to refrain from taking medications to treat or prevent their headaches.

### Standard Protocol Approvals, Registrations, and Patient Consents

Study participants provided written informed consent in accordance with each site’s Institutional Review Board guidelines. Sharing of anonymized data elements between sites was permitted via an institutional Data Transfer and Use Agreement.

### Clinical and Cognitive-Affective Measures

Participants completed a series of standardized self-report questionnaires to characterize clinical and affective symptoms including the Patient Health Questionnaire9 (PHQ-9)^29^ to assess depression, the Generalized Anxiety Disorder 7-item Scale (GAD-7)^30^ to assess anxiety, and the Pain Catastrophizing Scale (PCS)^31^ to assess negative cognitions and emotions during pain. Migraine disease characteristics, including headache frequency (days per month) and headache intensity (0-10 numerical rating scale) were also determined. To characterize group differences in clinical and demographic data, independent samples or Welch t-tests were used as appropriate using SPSS version 25. For categorical data, Chi-Square statistics (or Fisher’s exact test for smaller samples) were conducted.

### MRI Acquisition

Participants underwent a single 3T MRI scanning session whereby high-resolution structural and resting-state functional scans were acquired. Participants with CM were scanned on a GE MRI system (GE Healthcare, Milwaukee, WI) using an 8-channel phased array head coil. T1-weighted 3D axial FSPGR IR prep structural images (repetition time (TR)= 5.9 ms; echo time (TE)= minimum; flip angle= 15°; voxels= 0.9 × 0.9 mm; matrix size= 256 × 256; slice thickness = 1 mm, no gap) and T2*-sensitive gradient echo spiral-pulse functional images (TR= 2,000 ms; TE= 30 ms; flip angle= 80°; voxels= 3.4 × 3.4 mm; slice thickness= 4 mm; spacing= 0.5 mm) were obtained for volumetric and resting-state functional analyses, respectively. Participants with EM were scanned on a Tim Trio scanner with a 32-channel head coil. T1-weighted MPRAGE structural scans (repetition time (TR)= 2300 ms; echo time (TE)= 2.98 ms; flip angle= 9°; voxels= 1.0 × 1.0 mm; matrix size= 256 × 256; slice thickness = 1 mm, no gap) and T2-weighted echo-planar imaging functional scans (TR= 2,000 ms; TE= 28 ms; flip angle= 77°; voxels= 3.4 × 3.4 mm; slice thickness= 4 mm, no gap) were obtained.

### MRI Processing and Analysis

Amygdala volumes were quantified using a voxel-based morphometry approach implemented in FMRIB’s Software Library (FSL) v.5.0.1 (https://fsl.fmrib.ox.ac.uk/fsl/fslwiki/). Preprocessing of anatomical scans included the removal of non-brain tissues using the Brain Extraction Tool (BET)^32^ and tissue classification into gray matter, white matter, and cerebrospinal fluid using the Automated Segmentation Tool^33^. Scans were then aligned to standard space using the MNI152 2mm template atlas. To create a study-specific template, images were averaged and individual patient scans were non-linearly re-registered to the new template. A modulation step was carried out to scale gray matter by the amount of contraction or expansion that occurred during spatial normalization. Images were smoothed using a Gaussian kernel (4.6mm FWHM). To assess group differences in amygdala volume, a mask comprising the left and right amygdala was made using the Harvard-Oxford subcortical atlas (**Figure 1A**). A voxelwise general linear model was applied with age and sex as variables of no interest. Statistical significance was determined with permutation-based non-parametric testing using FSL’s randomise (thresholded at *p*< 0.05), and corrected for multiple comparisons using threshold-free cluster enhancement^34^, controlling the family-wise error rate. The Juelich histological atlas, which divides the amygdala into three regions (superficial/corticoid, basolateral, centromedial)^35^ (**Figure 1A**), was used to more precisely determine the location of amygdala volumetric differences. Relationships between amygdala volume, headache, and cognitive-affective measures were also examined using Pearson (r) or Spearman’s rho (r_s_) correlation coefficients in SPSS. To control for multiple comparisons, a Bonferroni correction was applied such that the p-value threshold for correlations was set to *p*< 0.005 (0.05/10) (five measures for each amygdala seed).

**Figure 1:**
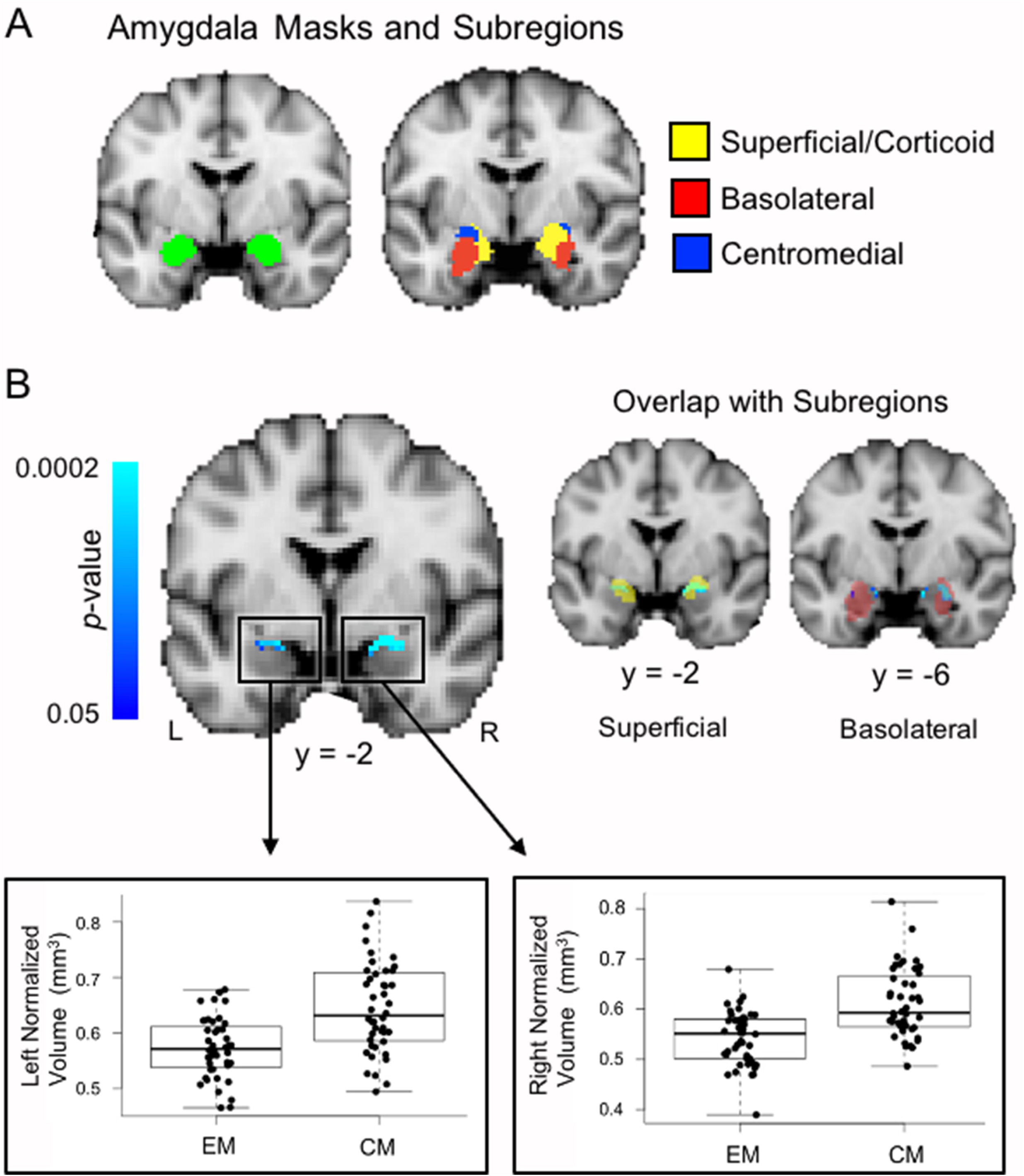
Voxel-based morphometry (VBM) assessment of amygdala volume. A VBM approach was used to assess amygdala volumes between groups. (A) A mask (green) comprised of both the left and the right amygdala was used to assess volume differences between patients with EM and CM (left). Amygdala subregions were classified according to labels provided by the Juelich histological atlas^35^ (right). (B) Compared to EM, patients with CM had significantly larger amygdala volumes bilaterally as indicated by the blue clusters. Boxplots and individual volume data are shown for each the left and right amygdala clusters (red boxes). To more precisely determine the location of significant group differences in volume, masks of amygdala subregions were overlay on top of the significant clusters. Only the superficial/corticoid and basolateral nuclei overlapped with group result as shown separately for each subregion (right). Amygdala masks and results presented in MNI 2mm standard space (per the analyses). Abbreviations: L: left; R: right; EM: episodic migraine; CM: chronic migraine.

Resting-state fMRI data were preprocessed using CONN functional connectivity toolbox v.18.b (https://www.nitrc.org/projects/conn)^36^, a Matlab-based software, in combination with Statistical Parametric Mapping (SPM12) (https://www.fil.ion.ucl.ac.uk/spm/). Preprocessing steps included slice time correction, functional realignment using b-spline interpolation, unwarping, co-registration of functional and structural scans, tissue segmentation of structural scans, normalization of structural and functional scans to the Montreal Neurological Institute (MNI) standard space, resampling to 2mm isotropic voxels, and spatial smoothing using an 8mm full-width at half maximum (FWHM) Gaussian smoothing kernel. Functional scans were band-pass filtered (0.008-0.09 Hz) and subject-specific covariates related to motion (six rotation/translation), white matter, and cerebrospinal fluid (using level one eroded masks to minimize partial volume effects with gray matter) were removed using a component-based noise correction approach (aCompCor)^37^. Additionally, volumes with framewise displacement (FD) larger than 0.5mm and global signal intensity changes greater than 3 standard deviations (default thresholds)^36^ as detected by Artifact Detection Tools (ART: https://www.nitrc.org/projects/artifact_detect/), were considered outliers and included as regressors in the first-level analysis. Quality control measures, including the examination of motion and functional connectivity (FC) associations, were taken at each level of preprocessing to ensure that results were not impacted by noise.

CONN toolbox was also used to assess amygdala connectivity and visualize functional brain data. Using a seed-based resting-state FC approach, subject-level correlation coefficients were calculated using the blood-oxygen-level-dependent (BOLD) signal time course extracted from each the left and right amygdala and nodes from three core large-scale cognitive brain networks: default mode (DMN), salience (SN), and central executive (CEN)^26,38^ (**Figure 2A**). These three networks were selected as they have been shown to be particularly important for understanding cognitive and affective function and dysfunction in psychiatric and neurologic disorders ^26^ and have previously been studied in patients with migraine^22–24,39^. Brain regions characterizing the DMN (medial prefrontal cortex, precuneus/posterior cingulate cortex, bilateral lateral parietal cortex), SN (anterior cingulate cortex, bilateral anterior insula, and rostral prefrontal cortex, and supramarginal gyrus (inferior part of Brodmann Area 40)), and CEN (bilateral lateral prefrontal cortex and posterior parietal cortex (posterior part of Brodmann Area 40)) were derived from CONN toolbox atlas labels (**Table 1**) based on independent component analyses of the Human Connectome Project (HCP) dataset of 497 subjects. For comparability across the literature, MNI coordinates of network labels were also provided in addition to anatomical functional network taxonomy labels, which have recently been proposed as a means towards developing a common resting-state nomenclature^40^ (**Table 1**). Amygdala seed regions were defined using the Harvard-Oxford Atlas. At the second level, amygdala connectivity was compared between groups controlling for age and sex by examining left and right amygdala connectivity to all network nodes. To examine progressive symptoms across all patients (EM and CM), associations between resting-state fMRI connectivity of each amygdala to all network nodes, headache severity (headache frequency and intensity), and cognitive-affective (depression, anxiety, pain catastrophizing) measures were separately evaluated using general linear models controlling for age and sex. Omnibus effects for group differences and associations with clinical and cognitive-affective measures assessed left and right amygdala connectivity to the nodes of all three networks using *F*-tests (degrees of freedom= (1,m), with 1 referring to the amygdala seed of interest and m referring to the number of subjects minus the rank of the design matrix) and follow-up *t*-tests revealed which connections were specifically contributing to group differences or associations using a two-sided false discovery rate (FDR)-corrected threshold of *q*< 0.05.

**Figure 2:**
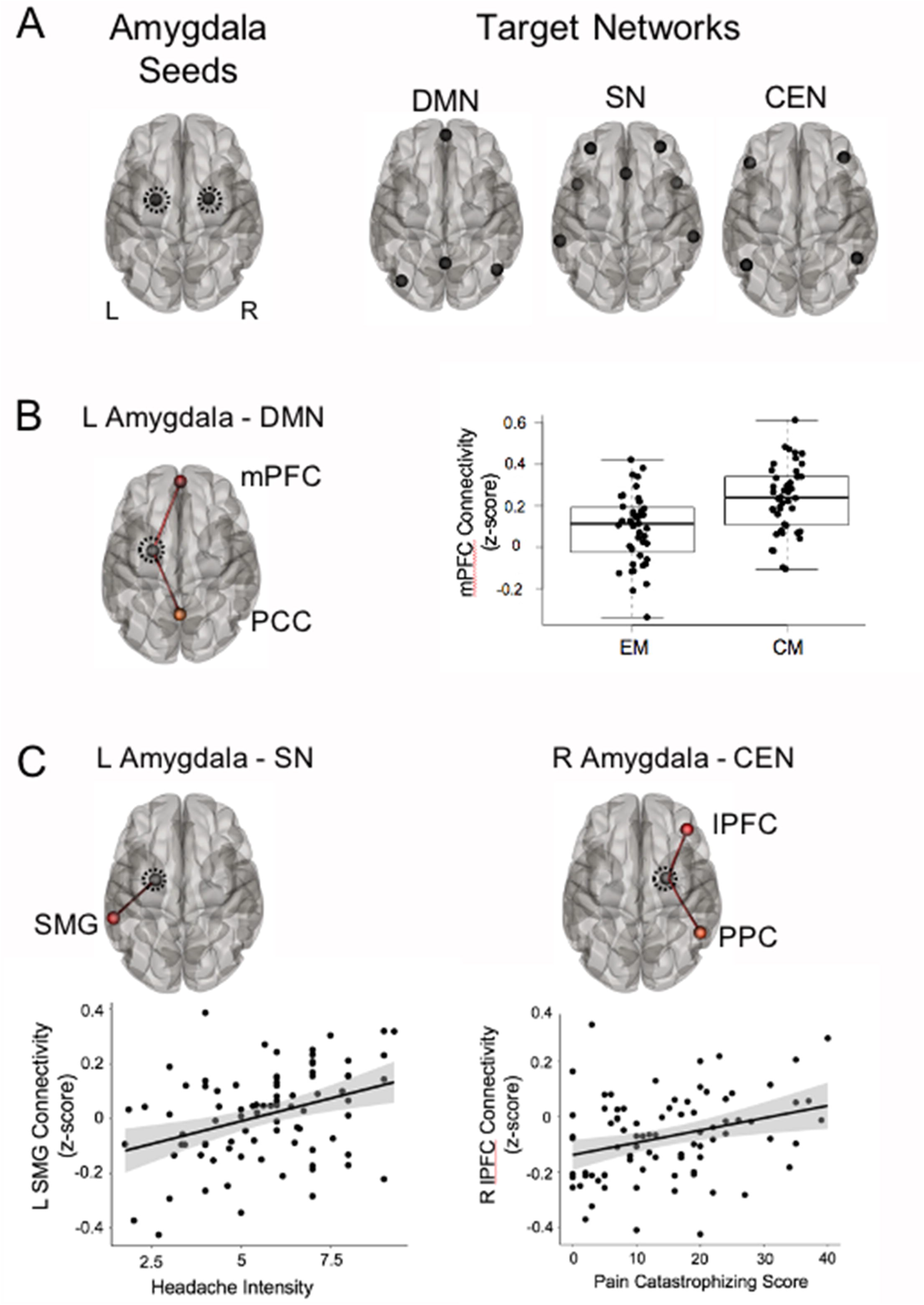
Amygdala functional connectivity with three core large-scale cognitive brain networks and associations with clinical/cognitive-affective measures. Resting-state functional connectivity analyses between the amygdala and three core large-scale brain networks were conducted to examine group differences between EM and CM and associations with clinical and affective measures. (A) Each the left and right amygdala were used as seeds to assess functional connectivity to target nodes within the DMN, SN, and CEN. (B) Compared to EM, patients with CM had increased connectivity between the left amygdala and the DMN (mPFC and PCC nodes). Boxplots for each group show the distribution of connectivity values to the mPFC node, with thicker black lines indicating median values. Individual connectivity values for each patient are also included. (C) Across all patients, left amygdala connectivity values to the SN (left SMG node) was positively associated with headache intensity ratings (left) and right amygdala connectivity to the CEN (lPFC and PPC nodes) was positively associated with pain catastrophizing scores (right). Shaded area on graphs represent 95% confidence intervals. Abbreviations: L: left; R: right; DMN: default mode network; SN: salience network; CEN: central executive network; mPFC: medial prefrontal cortex; PCC: precuneus/posterior cingulate cortex; SMG: supramarginal gyrus; lPFC: lateral prefrontal cortex; PPC: posterior parietal cortex.

**Table 1:**
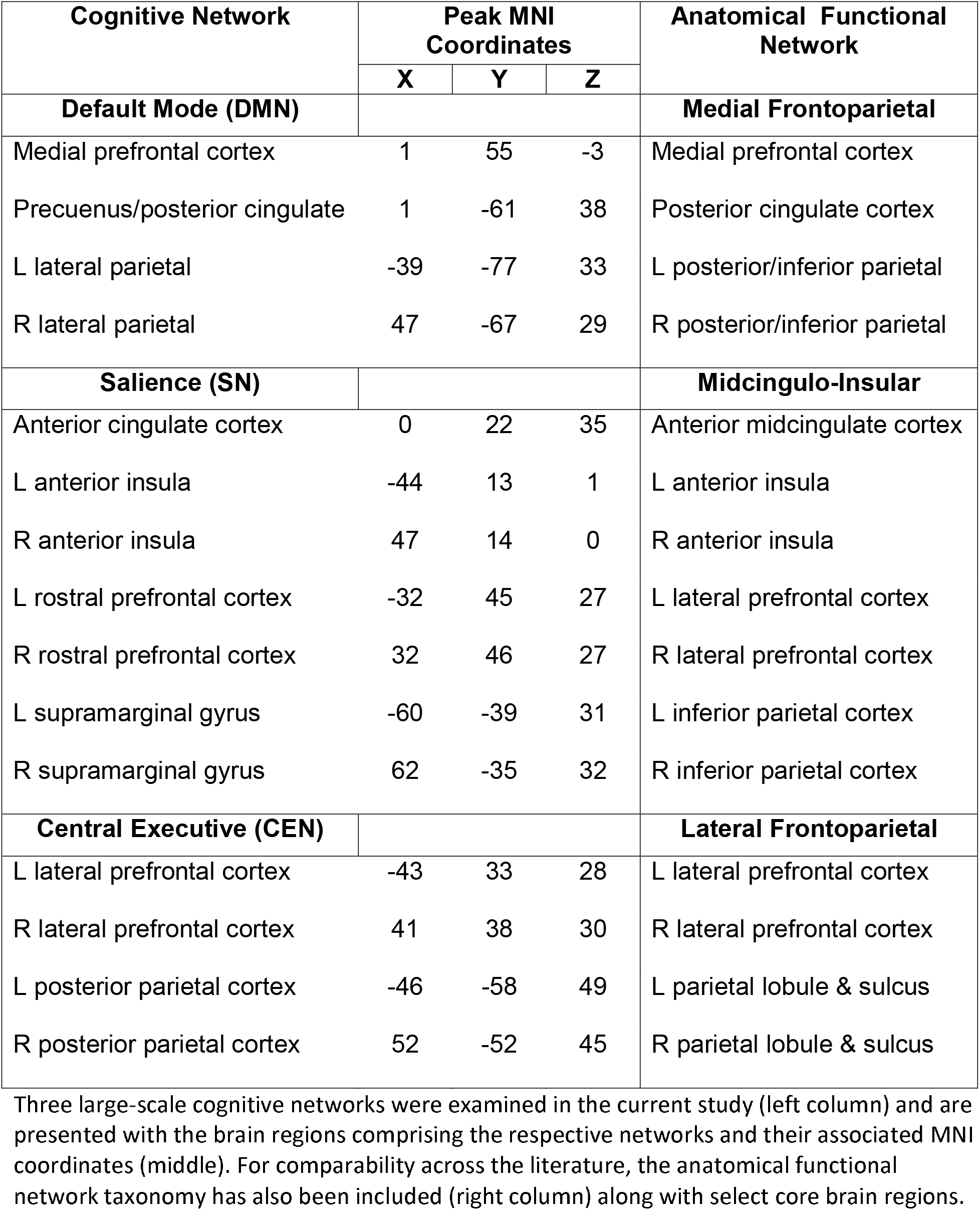
Resting-State Network Details.

| Cognitive Network | Peak MNI Coordinates |  |  | Anatomical Functional Network |
| --- | --- | --- | --- | --- |
|  | X | Y | Z |  |
| **Default Mode (DMN)** |  |  |  | **Medial Frontoparietal** |
| Medial prefrontal cortex | 1 | 55 | -3 | Medial prefrontal cortex |
| Precuneus/posterior cingulate | 1 | -61 | 38 | Posterior cingulate cortex |
| L lateral parietal | -39 | -77 | 33 | L posterior/inferior parietal |
| R lateral parietal | 47 | -67 | 29 | R posterior/inferior parietal |
| **Salience (SN)** |  |  |  | **Midcingulo-Insular** |
| Anterior cingulate cortex | 0 | 22 | 35 | Anterior midcingulate cortex |
| L anterior insula | -44 | 13 | 1 | L anterior insula |
| R anterior insula | 47 | 14 | 0 | R anterior insula |
| L rostral prefrontal cortex | -32 | 45 | 27 | L lateral prefrontal cortex |
| R rostral prefrontal cortex | 32 | 46 | 27 | R lateral prefrontal cortex |
| L supramarginal gyrus | -60 | -39 | 31 | L inferior parietal cortex |
| R supramarginal gyrus | 62 | -35 | 32 | R inferior parietal cortex |
| **Central Executive (CEN)** |  |  |  | **Lateral Frontoparietal** |
| L lateral prefrontal cortex | -43 | 33 | 28 | L lateral prefrontal cortex |
| R lateral prefrontal cortex | 41 | 38 | 30 | R lateral prefrontal cortex |
| L posterior parietal cortex | -46 | -58 | 49 | L parietal lobule & sulcus |
| R posterior parietal cortex | 52 | -52 | 45 | R parietal lobule & sulcus |

Three large-scale cognitive networks were examined in the current study (left column) and are presented with the brain regions comprising the respective networks and their associated MNI coordinates (middle). For comparability across the literature, the anatomical functional network taxonomy has also been included (right column) along with select core brain regions.

*<Insert Table 1>*

### Data Availability

Anonymized data can be shared upon request from qualified investigators.

## RESULTS

A total of 88 participants [44 with EM (36 women, 8 men; mean age ± SD: 37.8 ± 12.1) and 44 with CM (36 women, 8 men; mean age ± SD: 37.5 ± 12.1)] were included in the study. Patients with EM were selected to match patients with CM in terms of age and sex while maintaining equal numbers between groups. Clinical, demographic, and questionnaire details for each group are described in **Table 2**. In accordance with previous studies, patients with CM reported significantly higher levels of depression, anxiety, and pain catastrophizing compared to patients with EM^3,41^.

**Table 2:**
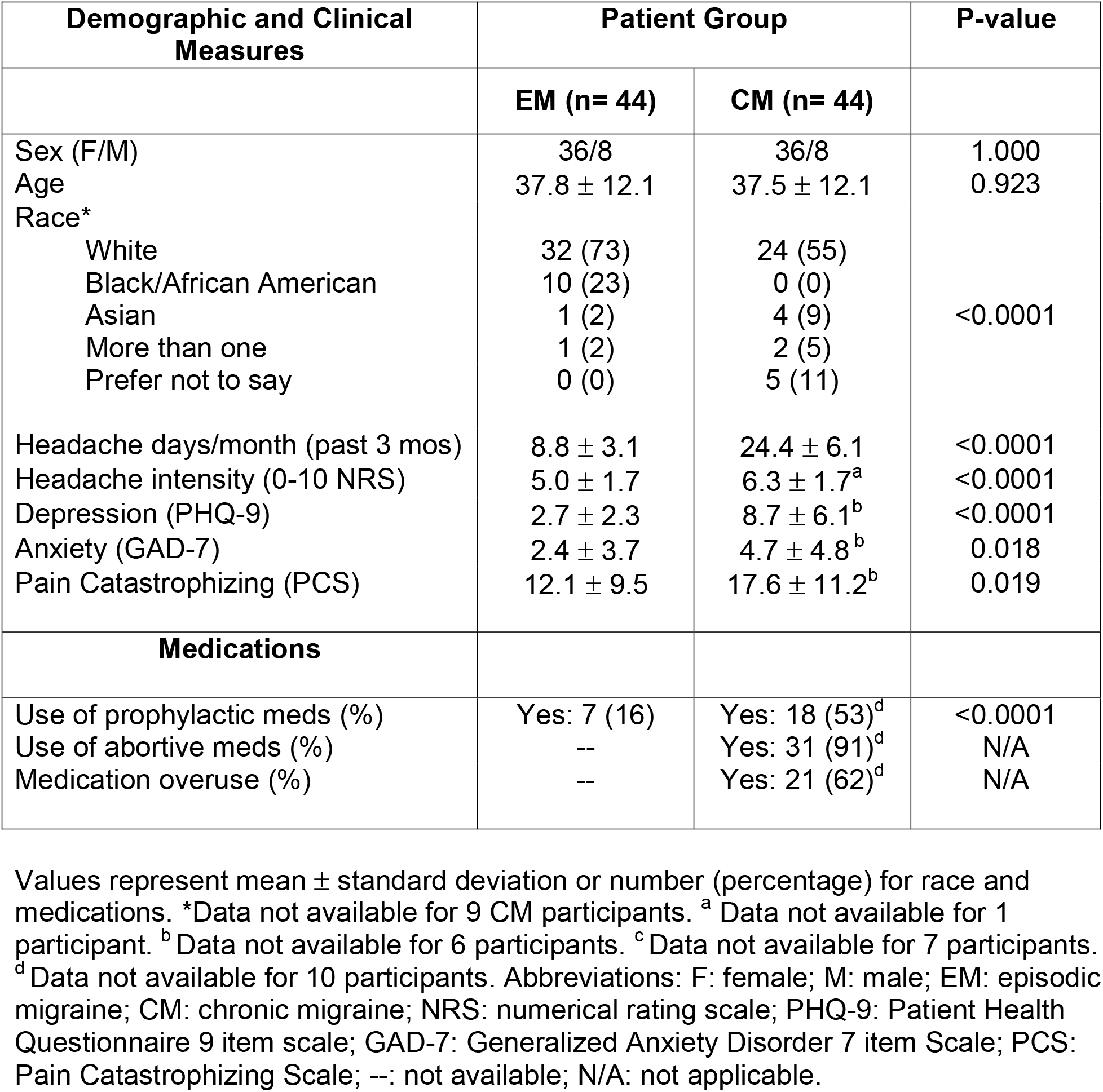
Summary of Patient Demographic and Clinical Features.

| Demographic and Clinical Measures | Patient Group |  | P-value |
| --- | --- | --- | --- |
|  | EM (n= 44) | CM (n= 44) |  |
| Sex (F/M) | 36/8 | 36/8 | 1.000 |
| Age | 37.8 ± 12.1 | 37.5 ± 12.1 | 0.923 |
| Race* |  |  |  |
| White | 32 (73) | 24 (55) | <0.0001 |
| Black/African American | 10 (23) | 0 (0) |  |
| Asian | 1 (2) | 4 (9) |  |
| More than one | 1 (2) | 2 (5) |  |
| Prefer not to say | 0 (0) | 5 (11) |  |
| Headache days/month (past 3 mos) | 8.8 ± 3.1 | 24.4 ± 6.1 | <0.0001 |
| Headache intensity (0-10 NRS) | 5.0 ± 1.7 | 6.3 ± 1.7<sup>a</sup> | <0.0001 |
| Depression (PHQ-9) | 2.7 ± 2.3 | 8.7 ± 6.1<sup>b</sup> | <0.0001 |
| Anxiety (GAD-7) | 2.4 ± 3.7 | 4.7 ± 4.8<sup>b</sup> | 0.018 |
| Pain Catastrophizing (PCS) | 12.1 ± 9.5 | 17.6 ± 11.2<sup>b</sup> | 0.019 |
| **Medications** |  |  |  |
| Use of prophylactic meds (%) | Yes: 7 (16) | Yes: 18 (53)<sup>d</sup> | <0.0001 |
| Use of abortive meds (%) | -- | Yes: 31 (91)<sup>d</sup> | N/A |
| Medication overuse (%) | -- | Yes: 21 (62)<sup>d</sup> | N/A |

Values represent mean ± standard deviation or number (percentage) for race and medications. \*Data not available for 9 CM participants. <sup>a</sup> Data not available for 1 participant. <sup>b</sup> Data not available for 6 participants. <sup>c</sup> Data not available for 7 participants. <sup>d</sup> Data not available for 10 participants. Abbreviations: F: female; M: male; EM: episodic migraine; CM: chronic migraine; NRS: numerical rating scale; PHQ-9: Patient Health Questionnaire 9 item scale; GAD-7: Generalized Anxiety Disorder 7 item Scale; PCS: Pain Catastrophizing Scale; --: not available; N/A: not applicable.

*<Insert Table 2>*

### Amygdala Volume Enlargement in CM

The regional VBM analysis of amygdala volumes revealed that compared to EM, patients with CM showed larger amygdala volumes bilaterally, spanning the basolateral (BLA) and superficial/corticoid nuclei as determined by the Juelich histological atlas (right cluster: 75 voxels, peak MNI coordinates: x= 26, y= -2, z= -14; left cluster: 30 voxels, peak MNI coordinates: x= -16, y= -4, z= -14) (**Figure 1B**). In examining associations with amygdala volumes derived from the group comparison, left amygdala volume was significantly correlated with headache frequency (r_s_= 0.34, *p*= 0.001) and headache intensity (r= 0.31, *p*= 0.003) (**Figure 3**). Right amygdala volume was correlated with headache frequency (r_s_= 0.42, *p*< 0.0001), headache intensity (r= 0.35, *p*= 0.001), and depression (r_s_= 0.35, *p*= 0.001). No other volumetric associations were significant according to the *p*< 0.005 Bonferroni-corrected threshold: right amygdala and PCS (r= 0.26, *p*= 0.026), right amygdala and GAD-7 (r_s_= 0.19, *p*= 0.084), left amygdala and depression (r_s_= 0.22, *p*= 0.052), left amygdala and PCS (r= 0.19, *p*= 0.075), and left amygdala and GAD-7 (r_s_= -0.028, *p*= 0.800).

**Figure 3:**
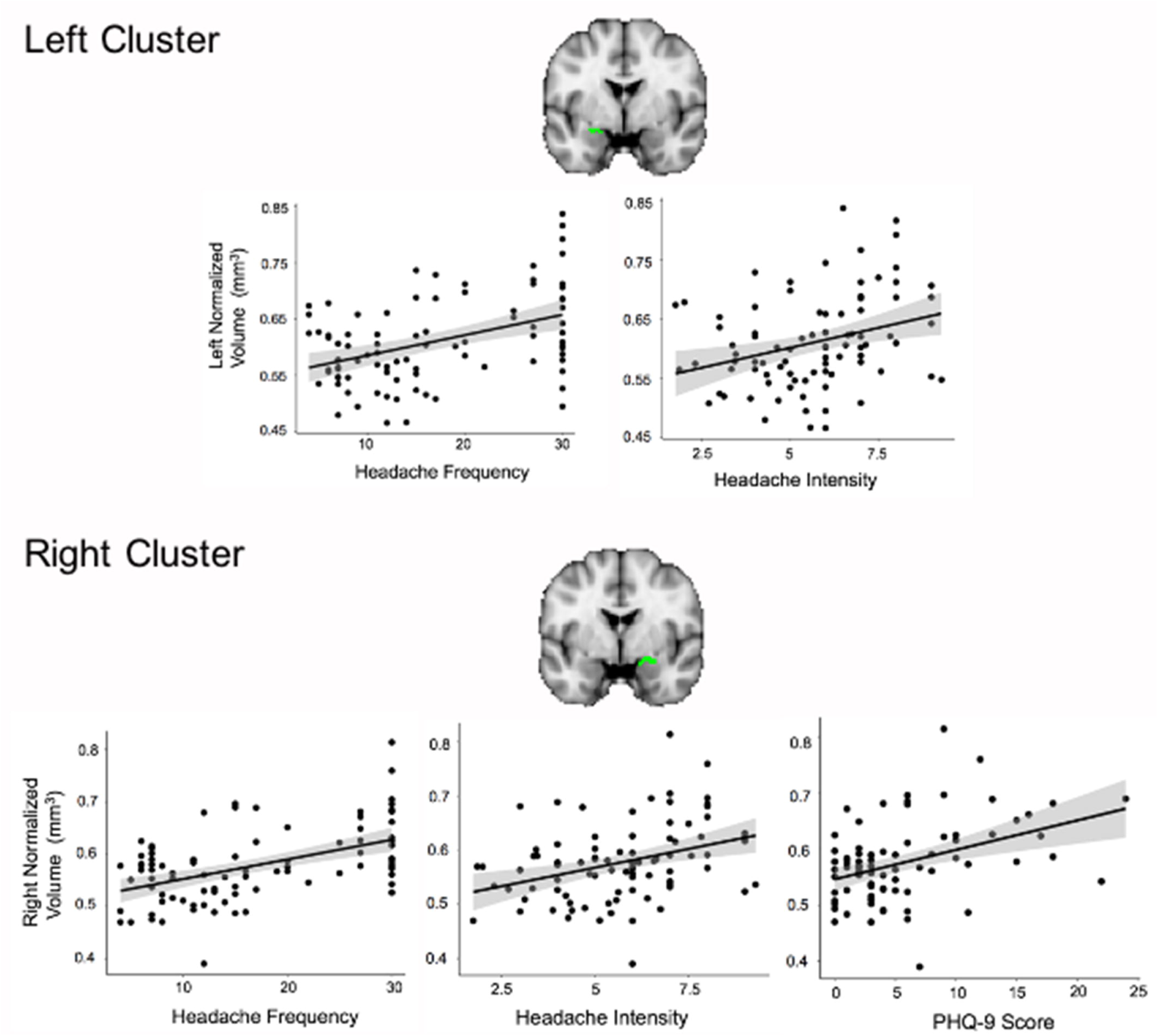
Significant Amygdala Volume Correlations. Relationships between amygdala volumes derived from the VBM group-contrast (cluster masks in green), headache severity, and cognitive-affective measures were assessed. Left amygdala volumes were positively associated with headache frequency (r_s_= 0.34) and headache intensity (r= 0.31) (top panel). Positive relationships were also observed between right amygdala volumes and headache frequency (r_s_= 0.42), headache intensity (r= 0.35), and depression (r_s_= 0.35) as measured by the PHQ-9 (bottom panel). Shaded area on graphs represent 95% confidence intervals. All correlations displayed were significant at the *p*< 0.005 level.

*<Insert Figure 1>*

### Increased Amygdala Functional Connectivity in CM

Amygdala connectivity to all network nodes was assessed between groups. Compared to EM, patients with CM showed greater left amygdala FC (*F*= 2.43, *p*= 0.007). Follow-up *t*-tests revealed that network nodes with increased FC to the left amygdala belonged to the DMN and included the medial prefrontal cortex (mPFC) (*t*= 3.96, *q*= 0.002) and precuneus/posterior cingulate cortex (PCC) (*t*= 3.38, *q*= 0.008) (**Figure 2B**). A follow-up analysis stemming from this group finding specifically assessed the relationship between left amygdala to DMN (mPFC and PCC) FC and clinical symptoms for all patients and revealed a positive relationship between left amygdala-mPFC FC and headache frequency (*t*= 2.51, *q*= 0.028).

*<Insert Figure 2>*

### Amygdala Functional Connectivity is Correlated with Progression of Headache and Cognitive-Affective Symptoms

To gain insight into the relationship between amygdala FC and the progression of clinical symptoms across all patients (EM and CM), we examined associations between amygdala FC to all network nodes and headache and cognitive-affective measures using a GLM approach. Left amygdala FC was associated with headache intensity (*F*= 3.17, *p*= 0.0006) (n= 86 due to missing questionnaire data and one participant having FC values > 3 standard deviations from the mean), with FC to the left supramarginal gyrus node of the SN driving the result (*t*= 4.14, *q*= 0.001) (**Figure 2C, left**). Additionally, right amygdala FC was associated with pain catastrophizing (*F*= 2.00, *p*= 0.029) (n= 82 due to missing questionnaire data) with t-tests revealing that nodes of the CEN (right lateral prefrontal cortex (*t*= 3.09, *q*= 0.038) and right posterior parietal cortex (*t*= 2.88, *q*= 0.038) were driving the result (**Figure 2C, right**). No relationships were observed between amygdala connectivity and anxiety scores (left amygdala: *F*= 0.59, *p*= 0.875; right amygdala: *F*= 0.77, *p*= 0.701).

*<Insert Figure 3>*

## DISCUSSION

Using a multimodal MRI approach, we showed that CM is associated with alterations in amygdala structure and large-scale cognitive network connectivity compared to EM. Our results indicate that patients with CM show larger amygdala volumes bilaterally and increased left amygdala-DMN FC compared to patients with EM. Moreover, positive associations were observed between: 1) left amygdala volume and headache frequency and intensity, 2) right amygdala volume and headache frequency, headache intensity, and depression, 3) left amygdala-DMN connectivity and headache frequency, 4) left amygdala-SN connectivity and headache intensity, and 5) right amygdala-CEN connectivity and pain catastrophizing. Taken together, our findings demonstrate structural and functional reorganization of the amygdala in CM compared to EM and support differential roles of the left and right amygdala in relation to the progression of headache and cognitive-affective symptoms in patients with migraine.

The amygdala is a highly interconnected limbic structure that plays a crucial role in the integration of sensory (including nociceptive) and emotional components of pain perception^9,10,42^. It has been extensively studied in the context of acute and persistent pain^9,10,13^, emotional processing^43^, and affective disorders including depression and anxiety^44,45^. Of relevance to migraine, the amygdala contains populations of calcitoningene-related peptide (CGRP)-expressing neurons^46^ and is one of the regions involved in the exacerbation of pain via anticipation, with clinical evidence suggesting this is particularly bothersome for disorders involving pain attacks^8,47^. The amygdala can be divided into three main groups based on cytoarchitectonic organization: 1) superficial/corticoid, 2) BLA, and 3) centromedial^35^. The superficial group is located adjacent to the BLA and is involved in olfactory and affective processing^48^. The BLA facilitates associative learning processes such as fear conditioning via afferents from cortical and subcortical regions including the thalamus, hippocampus, anterior cingulate cortex, and prefrontal cortex^49^. The BLA network is also involved in the attachment of emotional significance to broad sensory stimuli via associative learning^43,50^, which is particularly relevant since migraine is intrinsically a multisensory disorder with symptoms spanning the visual (photophobia), auditory (phonophobia), and olfactory (osmophobia) systems, among others^8^. The centromedial nuclei play a major role in generating behavioral responses via forebrain and brainstem projections involved in pain modulation.

As the progression of EM to CM involves disabling increases in sensory and affective symptoms^2,3^, examining the role of the amygdala in chronic migraine has become a growing research interest in the neuroimaging literature^16,17,51^. In one study investigating patients with CM, a region-of-interest approach was used to examine resting-state FC of five brain regions involved in affective processing (bilateral amygdala, bilateral anterior insula, and anterior cingulate cortex) to the whole brain^16^. Compared to healthy controls, patients with CM demonstrated less negatively correlated activity between the left amygdala and the superior frontal lobe and more negatively correlated activity between right amygdala and the left occipital lobe. Amygdala FC values were not associated with any of the clinical parameters investigated. In another study, Chen and colleagues examined amygdala volume and resting-state FC in patients with EM, CM, and healthy controls^17^. The results showed that patients with EM had increased FC of the left amygdala (primarily to the middle cingulate and precuneus cortex) compared to controls while patients with CM had increased amygdala FC (primarily to frontal, temporal regions) bilaterally compared to EM. Interestingly, only sleep quality was related to the FC findings and no statistically significant volume differences were reported. While the studies described here provide early evidence for altered amygdala connectivity in migraine chronification, noted study limitations include small sample sizes (≤ 20 participants per group), lack of an EM cohort and sex-matched controls in the first study, and lack of associations linking altered amygdala connectivity to sensory and/or cognitive-affective measures. The current study aimed to overcome these limitations by examining amygdala structure and FC in a larger cohort of EM and CM patients tightly matched for age and sex. Additionally, amygdala FC to three large-scale cognitive brain networks was specifically assessed as these networks provide powerful frameworks for investigating cognitive and affective functions across numerous psychiatric and neurologic disorders.

A major finding in the present study was the bilateral increase in amygdala volumes in patients with CM compared to EM spanning the BLA and superficial/corticoid nuclei. While the precise mechanisms underlying gray matter volume differences are not fully understood, candidate mechanisms include changes in dendritic branching/synaptogenesis, neurogenesis, axon sprouting, changes in glial cell number and/or morphology, and angiogensis^52^. Altered amygdala volumes have previously been reported in studies of migraine^21,53^, with one study demonstrating a positive relationship between left amygdala volume and headache frequency^53^. We replicate this finding and expand upon it by showing that left amygdala volumes were also positively associated with headache intensity. In a meta-analysis examining amygdala activation in patients with clinical pain, the most robust probability of activity was shown in left BLA (89%) (with additional probabilities noted in the superficial region) and a second cluster seated in the superficial regions of the right amygdala (68%)^10^. Clusters of significant volume differences between patient groups in the current study also spanned these amygdala subregions corroborating their potential role in migraine pain.

While measures of headache severity were positively associated with amygdala volumes bilaterally, only right amygdala volumes were associated with depression, suggesting a differential role for the left and right amygdala in the progression of clinical symptoms. Previous studies have shown that patients with CM are twice as likely to have depression compared to patients with EM and that depression is a risk factor for migraine chronification^3,4^. Aspects of depression related to negative valence (aversive and unpleasant) symptoms have been shown to be associated with amygdala function^54^. Recently, a distinct neural ensemble specifically in the BLA amygdala was shown to encode the negative affective valence of pain in mouse models of acute and chronic pain^12^. The right amygdala volume increase spanning the BLA in the current study may in part reflect the enhanced negative affective valence that occurs with migraine chronification.

Another major finding of our study was the significant increase in left amygdala connectivity to the DMN in patients with CM compared to EM. Resting-state FC describes the coordinated fluctuations of fMRI signals between brain regions at rest^55^. As such, patients with CM demonstrated a tighter coupling of brain fluctuations between the left amygdala and nodes of the DMN. The DMN is classically described as a task-negative network because its activity consistently decreases from baseline during a broad range of cognitive tasks, representing a “default mode of brain function”^56,57^. Over the years, the role of the DMN has grown to represent an integrated system for different aspects of self-referential thinking^26^. DMN connectivity has been studied across numerous neurologic and psychiatric disorders including chronic pain^58–62^; however, certain connectivity patterns appear to be disorder-specific. For example, enhanced DMN connectivity to the subgenual cingulate cortex is a recognized feature of major depressive disorder ^60,63^. Altered DMN connectivity has previously been reported in studies of migraine^22,23,39,64,65^, whereby EM or CM connectivity was compared to healthy controls. Specifically, within network connectivity of the DMN (DMN coherency) was reduced in patients with EM^64^ and in female patients with CM^23^. Additionally, reduced DMN connectivity to the CEN and enhanced DMN connectivity to the dorsal attention network was reported in patients with CM^22^. Other studies have shown DMN alterations that are related to clinical and/or cognitive-affective measures. For example, greater DMN to hypothalamus connectivity was reported in patients with CM compared to healthy controls; a finding which was negatively associated with migraine intensity^65^. In patients with EM, DMN to SN connectivity was positively associated with pain catastrophizing^39^. While the results of these studies varied based on the heterogeneity of imaging methods implemented and patient cohorts examined, it remains clear that migraine is associated with both intra- and inter-DMN network connectivity. We add to this literature by demonstrating that DMN connectivity to the left amygdala is enhanced in patients with CM compared to EM and that this connectivity is positively associated with headache frequency. Since the current study specifically compared patients with EM and CM as opposed to either patient cohort to controls and the group finding was related to how often headaches occur, our results are particularly relevant to the study of brain alterations associated with the progression of migraine symptoms. To our knowledge, only one other study directly assessed large-scale network connectivity in EM and CM^25^. Using a data-driven approach, Lee and colleagues examined group differences in network connectivity between migraine patient subtypes and healthy controls^25^. The results of their study indicated that compared to EM, patients with CM had greater connectivity in a network identified as the “pain matrix” because it included brain regions previously shown to involved in pain perception such as the dorsolateral prefrontal cortex, anterior insula, anterior cingulate cortex, thalamus, precuneus, and supramarginal gyrus, among other regions. The authors additionally examined the importance of each brain region comprising the identified pain network to determine which ones contributed most to the group result. The brain region with the greatest weight was identified as the anterior cingulate cortex, which overlapped with the medial prefrontal cortex, a node of the DMN. The second highest contributing brain region was the precuneus, which is also a DMN node. While using a different approach from the current study, altered connectivity of DMN nodes was also shown to best differentiate patients with EM from CM.

We also observed correlations between amygdala connectivity and clinical measures related to headache severity and cognitive-affective measures. Specifically, we showed that left amygdala connectivity to the SN was positively associated with headache intensity. The SN is a network involved in attention, interoceptive and affective processes^66^. It plays a key role in identifying the most relevant information from both internal and external stimuli to guide attention^66,67^. In our study, the tighter coupling of amygdala to SN connectivity with increasing headache intensity may reflect a hypervigilance or enhanced attention to headache pain. Migraine has been described as a disorder of “multisensory integration”^68^, whereby significance is assigned to nonthreatening sensory stimuli. The amygdala to SN connectivity increase in CM may reflect the abnormal assignment of importance to sensory perceptions that occur with migraine. This finding in line with the growing pain literature describing aberrant functioning of brain networks that assign salience values to stimuli in other chronic pain disorders^69^. Intranetwork connectivity of the SN has previously shown to be decreased in women with CM compared to controls^24^. Interestingly, a subgroup analysis also determined that precise SN connectivity patterns differed between patients with CM and CM with medication overuse headache. Since our CM group included patients with and without medication overuse headache, future studies aimed at investigating the relationship between amygdala to SN connectivity in additional migraine subgroups are warranted.

We also showed that right amygdala connectivity to the CEN was positively associated with pain catastrophizing. The CEN is a frontoparietal system anchored in the lateral prefrontal cortex and posterior parietal cortex. This network is important for working memory, rule-based problem solving, decision making, and goal-directed behaviors^26^. Recent studies of migraine have shown that altered CEN connectivity may be related to clinical features of migraine. For example, Coppola and colleagues reported that patients with CM have reduced CEN connectivity compared to controls^22^ and that lower CEN connectivity was associated with greater headache severity. In the current study, we show that enhanced connectivity of the CEN to the right amygdala is associated with higher pain catastrophizing; a construct describing maladaptive responses to pain-related distress^31^. As this association was previously reported in patients with chronic low back^14^, it may reflect maladaptive cognitive-affective changes that occur with pain chronification generally. Interestingly, stronger amygdala to posterior parietal cortex activity was recently reported in healthy individuals who demonstrate greater pain facilitation by negative emotions^70^. Since high levels of pain facilitation can be maladaptive in the long-term, future studies are needed to determine the degree to which this enhanced connectivity is predictive of the development of chronic pain disorders including CM.

There are some important study limitations to be considered when interpreting the results of the current study. First, MRI scans were acquired on different scanners for patients with EM and CM and precise MRI protocols differed between sites. It is possible that group differences in amygdala volumes or FC could in part be due to noise introduced by differences in scanning protocols. While we cannot completely rule this out, differences between EM and CM were significantly associated with clinical and/or cognitive-affective measures suggesting clinically meaningful findings. Another study limitation is that we examined a naturalistic cohort of patients with migraine, meaning patients were not asked to refrain from medication use and some patients with CM also had medication overuse headache. The precise effects of medications on amygdala structure and function were not assessed and could have impacted the findings. Lastly, our study was cross-sectional. While we have discussed implications for migraine progression by comparing patients with EM and CM and examining associations across all patients, longitudinal studies assessing changes in amygdala structure and function over time would be highly valuable and provide further insight in this domain.

The clinical management of migraine is challenging particularly when patients progress to CM. As such, there is a major need to understand the mechanisms underlying CM and migraine chronification. Toward this effort, we provide novel insight into how increased amygdala volume and FC to large-scale networks differ between patients with EM and CM and support a role for the amygdala in migraine symptom progression. These findings may inform comprehensive therapeutic strategies aimed at slowing or reversing migraine progression by reducing both headache and negative cognitive-affective symptoms.

## Data Availability

Anonymized data can be shared upon request from qualified investigators.

## ACKNOWLEDGEMENTS

We thank the study participants for their time and dedication to this research. We also thank Daniel A. Bissell for technical assistance with data organization. This work was supported by the SunStar Foundation to R.P. Cowan and NCCIH/NIH R01 AT007176 to D.A. Seminowicz.

## COMPETING INTERESTS

No competing interests for DDD, SRK, BMS, AP, VM, and DAS. RPC is an advisor (unrelated to current study) for Alder, Amgen, Allergan, Biohaven, Curex, Teva, and Xoc.

## Notes

### Clinical Trial

NCT03231241

### Author Declarations

Both Stanford University and the University of Maryland, Baltimore received institutional IRB approval. Data sharing between institutions was permitted via a Data Transfer and Use Agreement (AGR760476).

